# The neurodevelopmental spectrum: phenotypic architecture, etiology, predictive utility, and specificity across development

**DOI:** 10.1101/2025.07.31.25332492

**Authors:** Giorgia Michelini, Wangjingyi Liao, Shiqi Lu, Chiara Caserini, Thalia C. Eley, Angelica Ronald, Sylia Wilson, Margherita Malanchini, Kaili Rimfeld

**Author notes:** Correspondence to Dr Giorgia Michelini, G. E. Fogg Building, Queen Mary University of London, London, UK.

## Abstract

Neurodevelopmental conditions are highly heritable, heterogeneous, and frequently co-occur. Transdiagnostic dimensional approaches have advanced understanding and classification of psychiatric disorders, but have largely omitted neurodevelopmental conditions. Using longitudinal data for >10,000 children from the Twins Early Development Study, we investigated the structure of a transdiagnostic “neurodevelopmental spectrum” across development, its etiology, its ability to predict functional outcomes, and the specificity of these associations. Hierarchical exploratory factor modeling of a broad set of traits/symptoms delineated a neurodevelopmental spectrum encompassing neurodevelopmental traits at ages 7, 12, and 16. This spectrum emerged as a distinct dimension alongside separable dimensions of internalizing, externalizing, and psychosis symptoms. The neurodevelopmental spectrum was highly heritable across development in twin analyses (h^2^=0.60-0.82) and predicted by polygenic scores (PGS) for neurodevelopmental, cognitive, and educational phenotypes (R^2^ up to 2.30% in single-PGS analyses, 3.36% in multi-PGS analyses). Perinatal and early developmental factors (e.g., low birth weight, language delays) were also associated with this spectrum (R^2^ up to 8.65%). Individual differences in the neurodevelopmental spectrum predicted cognitive and educational outcomes both concurrently and longitudinally (R^2^ up to 20.61%), largely due to overlapping genetic effects. Associations with predictors and outcomes remained largely unchanged after adjusting for internalizing, externalizing, and psychosis dimensions, indicating they were specific to the neurodevelopmental spectrum and not attributable to shared variance with other co-occurring symptoms. Our novel results on the phenotypic architecture, etiological validity, predictive utility, and specificity of the neurodevelopmental spectrum across development support its integration into transdiagnostic frameworks, with important implications for advancing research, psychiatric classification, and clinical care.

## Introduction

Neurodevelopmental difficulties with attention, social communication, learning, cognition, movement, and language are heritable and early-emerging traits that characterize approximately 15% of children and adults worldwide, with many more showing subthreshold manifestations [1–3]. In traditional diagnostic manuals (i.e., DSM-5 and ICD-11), these difficulties are the defining features of “neurodevelopmental disorders”, including attention-deficit/hyperactivity disorder (ADHD), autism spectrum disorder, intellectual and specific learning disabilities, motor disorders (e.g., Tourette, developmental coordination disorder), and speech, language, and communication disorders [4, 5]. Despite being classified as distinct categories, neurodevelopmental conditions significantly overlap in their presentations and underpinnings [6–10]. They also co-occur with one another and with other psychiatric disorders in the majority of cases [11–14], either concurrently or at different points during development. The emphasis on individual diagnoses often leads to co-occurring conditions being overlooked and hence not addressed clinically (a phenomenon known as “diagnostic overshadowing”), contributing to worse clinical and functional outcomes [15–17]. New research and evidence-based clinical approaches are urgently needed to improve understanding, identification, prediction of future outcomes, and provision of accommodations and treatment/support strategies for neurodevelopmental and co-occurring conditions, in line with the priorities of the global neurodivergent community [15, 18].

Transdiagnostic psychiatric frameworks, including the Hierarchical Taxonomy of Psychopathology (HiTOP) [19], hierarchical causal taxonomies [20], and the Research Domain Criteria (RDoC) [21], are emerging as useful alternatives to categorical systems for understanding mechanisms and improving clinical care for individuals with psychiatric conditions [19–22]. The focus on dimensions (or “spectra”) addresses key limitations of categorical diagnoses, such as uncertain diagnostic boundaries and within-disorder heterogeneity [19, 23]. Moreover, by focusing on transdiagnostic dimensions that span multiple conditions, transdiagnostic frameworks offer a useful approach for assessing and accounting for widespread psychiatric co-occurrence [21, 22]. Examples of clinical dimensions in these frameworks include the general psychopathology (“p”) factor [24, 25] and broad externalizing, internalizing, and psychosis/thought disorder spectra [26], capturing dimensional features of many psychiatric conditions. Researchers and clinicians have recently highlighted the advantages of applying a transdiagnostic approach to neurodevelopmental conditions (e.g., the Early Symptomatic Syndromes Eliciting Neurodevelopmental Clinical Examinations [ESSENCE] framework) [6, 27–30], and its alignment with the priorities of the neurodivergent community [31, 32]. Yet, most of the literature that informed extant transdiagnostic psychiatric frameworks has not included neurodevelopmental conditions [19, 20, 24], despite their established co-occurrence with psychiatric conditions included in these frameworks [7]. Studies have also predominantly focused on adult samples [19, 33, 34], where neurodevelopmental traits are not commonly assessed. Consequently, current transdiagnostic frameworks omit the majority of neurodevelopmental conditions.

To overcome these limitations, we recently proposed the inclusion of a new “neurodevelopmental spectrum” into transdiagnostic psychiatric frameworks, bridging the emerging transdiagnostic focus on neurodevelopmental conditions with more established transdiagnostic approaches to other psychiatric conditions [7]. Building on a comprehensive evaluation of the literature, we defined this novel spectrum as a broad latent dimension reflecting the overlapping features of neurodevelopmental conditions, their similar developmental profiles, and their shared genetic, cognitive, and neural underpinnings. Transdiagnostic dimensional phenotypes are increasingly recognized as promising targets for advancing psychiatric nosology, uncovering mechanisms, and personalizing care [35–38]. Thus, conceptualizing neurodevelopmental traits in this spectrum and integrating them with psychiatric symptoms already included in transdiagnostic psychiatric frameworks can significantly advance research and clinical care for neurodevelopmental conditions and their common psychiatric comorbidities. However, key questions (Q) central to establishing this spectrum remain unanswered [7].

First, which features of neurodevelopmental conditions belong to the neurodevelopmental spectrum, as opposed to better-established transdiagnostic spectra (e.g., externalizing, internalizing, and psychosis/thought disorder spectra), across childhood and adolescence (Q1)? Available studies of the phenotypic architecture of neurodevelopmental and other psychiatric traits have included a limited set of neurodevelopmental features (usually ADHD and, to a lesser extent, autism) [7, 39]. They have also typically relied on cross-sectional data spanning wide age ranges [7, 27, 40]. As a result, the components and boundaries of the neurodevelopmental spectrum across development remain poorly defined. Clarifying these boundaries is important not only for understanding and classifying neurodevelopmental conditions, but also for informing models of other psychiatric conditions. For example, although psychotic disorders have been proposed to have “neurodevelopmental” origins [41, 42], no study has formally tested whether psychotic experiences cluster within a neurodevelopmental spectrum in children and young people. Second, what is the etiology of the neurodevelopmental spectrum across development (Q2)? Recent genomic structural equation modelling (gSEM) studies have identified genomic neurodevelopmental factors that mirror the phenotypic neurodevelopmental spectrum [43– 45]. However, further research is needed to establish its etiology―an important source of external validity evidence [41, 46]―particularly with respect to the joint contribution of genetic and environmental influences. Third, does the neurodevelopmental spectrum predict important outcomes across development (Q3)? Previous cross-sectional studies have limited the ability to test whether it predicts key outcomes (e.g., cognitive and academic functioning) over time, which is an essential step for evaluating its practical and clinical utility [47, 48]. Finally, are these associations with predictors and outcomes specific to the neurodevelopmental spectrum or shared with transdiagnostic dimensions capturing commonly co-occurring internalizing, externalizing, and psychosis symptoms (Q4)? Addressing this question is critical for determining whether the neurodevelopmental spectrum has distinct underpinnings and independently predicts outcomes beyond broader transdiagnostic effects.

Here, we addressed these complementary questions using longitudinal, multi-informant data collected across childhood and adolescence in a large, nationally representative study from the UK, the Twins Early Development Study (TEDS) [49, 50]. A deeper understanding of the phenotypic architecture (Q1), validity (Q2), utility (Q3), and specificity (Q4) of the neurodevelopmental spectrum across development promises to guide future psychiatric classification, genomic research, and precision/personalized medicine strategies. To address Q1, we examined the phenotypic architecture of a putative neurodevelopmental spectrum alongside better-established transdiagnostic spectra, by modeling a comprehensive set of neurodevelopmental and psychiatric symptoms/traits assessed in childhood and adolescence (ages 7, 12, and 16). To address Q2, we investigated the etiology of the neurodevelopmental spectrum across development using twin analyses, as well as associations with polygenic scores and perinatal/early developmental predictors previously linked to individual neurodevelopmental conditions, thereby providing evidence for the validity of this spectrum [27, 51, 52]. Turning to Q3, we examined phenotypic and genetic associations with key cognitive and educational outcomes, both cross-sectionally and longitudinally, to evaluate the predictive utility of this spectrum. Finally, we addressed Q4 by testing the specificity of the associations identified with predictors (Q2) and outcomes (Q3), with analyses examining whether associations remained after accounting for other transdiagnostic spectra (e.g., internalizing, externalizing). We hypothesized that the neurodevelopmental spectrum would capture neurodevelopmental traits across childhood and adolescence, display substantial heritability and associations with relevant genetic and perinatal/early developmental factors, and predict cognitive and educational outcomes across development, beyond associations with other transdiagnostic spectra.

## Methods

We pre-registered our research aims, hypotheses, and analyses on the Open Science Framework (https://osf.io/dqbnj/?view_only=f2510b6766bc4b86bf9344fee1139233) before accessing the data. For small deviations from the pre-registration, see Supplementary Methods 1.

### Participants

Our sample included participants from TEDS, a nationally representative longitudinal study of over 16,000 twin pairs (33% monozygotic, 67% dizygotic) born in England or Wales between 1994-1996 [49, 50]. Detailed information on recruitment, demographic characteristics, study procedures, ethical approvals, and sample representativeness is available elsewhere [49, 50]. The current study focused on participants with data available at age 7 (N=15,668, 51% female, mean age (SD)=7.07 (0.25)), 12 (N=12,465, 52% female, mean age (SD)=11.31 (0.71)), and 16 (N=10,261, 55% female, mean age (SD)=16.32 (0.68)). The socio-demographic characteristics of our study sample are well aligned with those of the whole cohort at first contact (when participants were approximately 18 months old) and the UK population at the time of recruitment [50]. For example, for participants completing age 7-16 assessments, 93% were White (vs. 92% at first contact and 93% national equivalent), 46% of mothers and 93% of fathers were employed (vs. 46% at first contact and 50% national equivalent for mothers; 93% at first contact and 91% national equivalent for fathers); and 40% of mothers and 48% of fathers completed A-level (i.e., high school) or higher degree (vs. 40% at first contact and 35% national equivalent for mothers; 47% at first contact and national equivalent for fathers) [50]. As TEDS also collected DNA in part of the sample, genomic data were available for a subset of participants included at age 7 (N=8,262), 12 (N=7,080), and 16 (N=6,030).

### Measures and procedures

#### Neurodevelopmental and psychiatric traits

At each wave, TEDS families completed broad assessments of child neurodevelopmental traits (ADHD and autism at ages 7, 12 and 16; learning, motor, and speech difficulties at age 7), internalizing problems, externalizing behaviors, psychotic experiences (age 16 only), and eating problems (age 16 only) [49, 50, 53]. Within each wave, measures were selected to reflect developmentally-relevant constructs at each developmental stage, meaning that measures partly overlapped across waves [49, 50]. Parent-reports were collected across waves, with additional self-reports collected at age 16. A detailed list of measures available at each age is available in Supplementary Table 1.

#### Polygenic scores

Procedures involving DNA collection, genotyping, quality control, and constructing polygenic scores have been described previously [54] and are available in Supplementary Methods 2. Polygenic scores were derived from the latest available genome-wide association studies (GWAS). In primary analyses, we included polygenic scores capturing genetic liability for neurodevelopmental conditions (i.e., ADHD, autism, Tourette) and cognitive, educational, and brain phenotypes, which we hypothesized to be most relevant to capturing the genetic underpinnings of the neurodevelopmental spectrum [55– 63] (Supplementary Table 2). Additional polygenic scores for other psychiatric disorders and personality traits were used in exploratory analyses.

#### Perinatal and early developmental predictors

We included key perinatal/early developmental predictors assessed at first contact, when participants were ∼18 months (i.e., birth weight and length, gestational age, physical problems in the first 18 months) [51] and during age 3 assessments (e.g., measures of developmental delays in the acquisition of cognitive and language abilities) previously associated with individual neurodevelopmental conditions [7]. For details, see Supplementary Table 3.

#### Cognitive and educational outcomes

Cognitive outcomes included measures of general cognitive ability (‘g’) at ages 7, 12, and 16, reading ability at ages 7 and 12, and reading comprehension and fluency at age 16 [64, 65]. Educational outcomes included teacher grades for core subjects (English, Mathematics, and Science) at ages 7 and 12 and standardized exam scores at age 16, averaged across subjects; parent-reported information about the longest time away from school, and teacher-reported behavior, learning, happiness, and effort in school relative to other students at age 7; teacher-reported information about referral to special education support at age 7 and receipt of support due to having special education needs at age 12. See Supplementary Table 4 for details.

### Statistical analyses

Primary analyses focused on data from neurodevelopmental and psychiatric assessments completed by parents, while self-reports were used in secondary analyses. Parents are reliable reporters of neurodevelopmental and co-occurring traits/symptoms across childhood and adolescence [66]. TEDS parents (mainly mothers) also completed the largest number of relevant assessments from age 7 to 16.

#### Phenotypic architecture of the neurodevelopmental spectrum (Q1)

To examine the joint phenotypic architecture of neurodevelopmental traits and psychiatric symptoms, we ran exploratory structural equation modeling (ESEM, an SEM-based exploratory factor analysis approach) in Mplus 8 [67] and the MplusAutomation R package [68]. Analyses were run on a total of 75, 104, and 130 parent-reported items covering neurodevelopmental and psychiatric traits available at ages 7, 12, and 16, respectively. Secondary analyses on self-reports at age 16 included 180 items. Items had skewed distributions and were specified as categorical, using the WLSMV estimator. Nesting of twins within families was accounted for using “CLUSTER” and “TYPE=COMPLEX” commands. We extracted factor solutions with an increasing number of factors, following established analytic procedures [27, 69, 70]. An exploratory approach was preferred over confirmatory factor analysis as the number of factors and their composition across hierarchical levels were uncertain, due to the small number of previous studies with relevant indicators. We also chose to run analyses in the whole sample (rather than running exploratory and confirmatory factor analyses on two subsets) to maximize statistical power. The maximum number of factors was determined with parallel analyses, with extraction stopped when eigenvalues fell within the 95% confidence interval of eigenvalues from simulated data [71], and based on the interpretability of factor solutions, defined as at least 4 primary loadings (highest loading ≥0.35 and at least 0.10 greater than all other loadings) for each factor [72, 73]. Factors were rotated using an oblique rotation (goemin) to allow the extracted factors to correlate, as expected [27].

To map the hierarchical structure and transitions between factor solutions, random-intercept mixed models (i.e., multi-level linear regressions clustering within the family to account for twin relatedness) [74] measured the association between factor scores from models with an increasing number of factors, using Goldberg’s bass-ackwards approach [75]. This approach was chosen as it is the only available method to delineate a hierarchical structure with 3+ levels through exploratory factor modeling. Factors in the derived hierarchical structure can be interpreted as interconnected across hierarchical levels, from a 1-factor level to more complex solutions with specific factors, allowing the examination of shift and reorganization of content across levels. As our hypotheses concerned a broad neurodevelopmental factor, we identified at each age the highest level of the hierarchy with a neurodevelopmental factor capturing traits of multiple neurodevelopmental conditions, and used these factors in subsequent analyses.

Random-intercept mixed models, accounting for family relatedness, age, and sex examined the phenotypic stability of neurodevelopmental factors at ages 7, 12, and 16, and the association between factors based on parent- and self-reports in secondary analyses.

#### Etiological validity (Q2): twin analyses, polygenic scores, and perinatal/early developmental predictors

The contribution of genetic and environmental effects on the neurodevelopmental factor at each age was quantified with the classic twin design (Supplementary Methods 3), controlling for age and sex [76]. Multivariate twin modeling estimated the overlap in genetic effects between neurodevelopmental factors over time.

We then ran random-intercept mixed models accounting for family relatedness to examine the association between polygenic scores and the neurodevelopmental factor across development. Analyses used the standardized residuals of the top 10 genetic principal components of population structure, genotyping chip, genotyping batch, age, and sex to control for these variables. Primary analyses focused on polygenic scores for neurodevelopmental conditions, cognitive and educational phenotypes, and brain measures [55–63] (Supplementary Table 2). In a pre-registered exploratory analysis, we further compared R^2^ in the neurodevelopmental factor explained by the most predictive polygenic score, R^2^ jointly explained by polygenic scores in primary analyses, and R^2^ jointly explained by a wider set of polygenic scores (Supplementary Table 2), using elastic net models with cross-validation (Supplementary Methods 4) [65, 77, 78].

Random-intercept mixed models also tested associations between perinatal/early developmental predictors and neurodevelopmental factors, accounting for family relatedness, age, and sex.

#### Predictive utility (Q3): Associations with cognitive and educational outcomes

We tested the cross-sectional association between the neurodevelopmental factors and cognitive and educational outcomes at each wave. Longitudinal models tested the association between the age-7 neurodevelopmental factor and age-12/16 cognitive and educational outcomes, and between the age-12 neurodevelopmental factor and age-16 cognitive and educational outcomes. Random-intercept mixed models were run in analyses of continuous outcomes (e.g., reading ability) and generalized estimating equations (GEE) in analyses of categorical outcomes (e.g., referral for special education support). The genetic and environmental effects underlying these associations were examined with bivariate twin models (Supplementary Methods 3).

#### Specificity of associations with predictors and outcomes (Q4)

To determine the specificity of the identified associations, we repeated the random-intercept mixed models addressing Q2 and Q3 at each wave also controlling for other transdiagnostic factors (e.g., internalizing, externalizing) in the same factor solution at which we extracted the neurodevelopmental factor.

## Results

### Neurodevelopmental spectrum across childhood and adolescence (Q1)

Modeling of parent-reported items delineated a hierarchical structure with a general (p) factor at the top and up to seven factors at age 7, six factors at age 12, and eight factors at age 16 (Figure 1; factor loadings and correlations are reported in Supplementary Tables 5-7). We found a clear factor capturing multiple neurodevelopmental traits across ages, first emerging in the 3-factor solution at age 7 and 12 and in the 5-factor solution at age 16. For full results (including factor loading and correlations for each factor solution), see Supplementary Results 1-3 and Supplementary Tables 5-7.

**Fig. 1.**
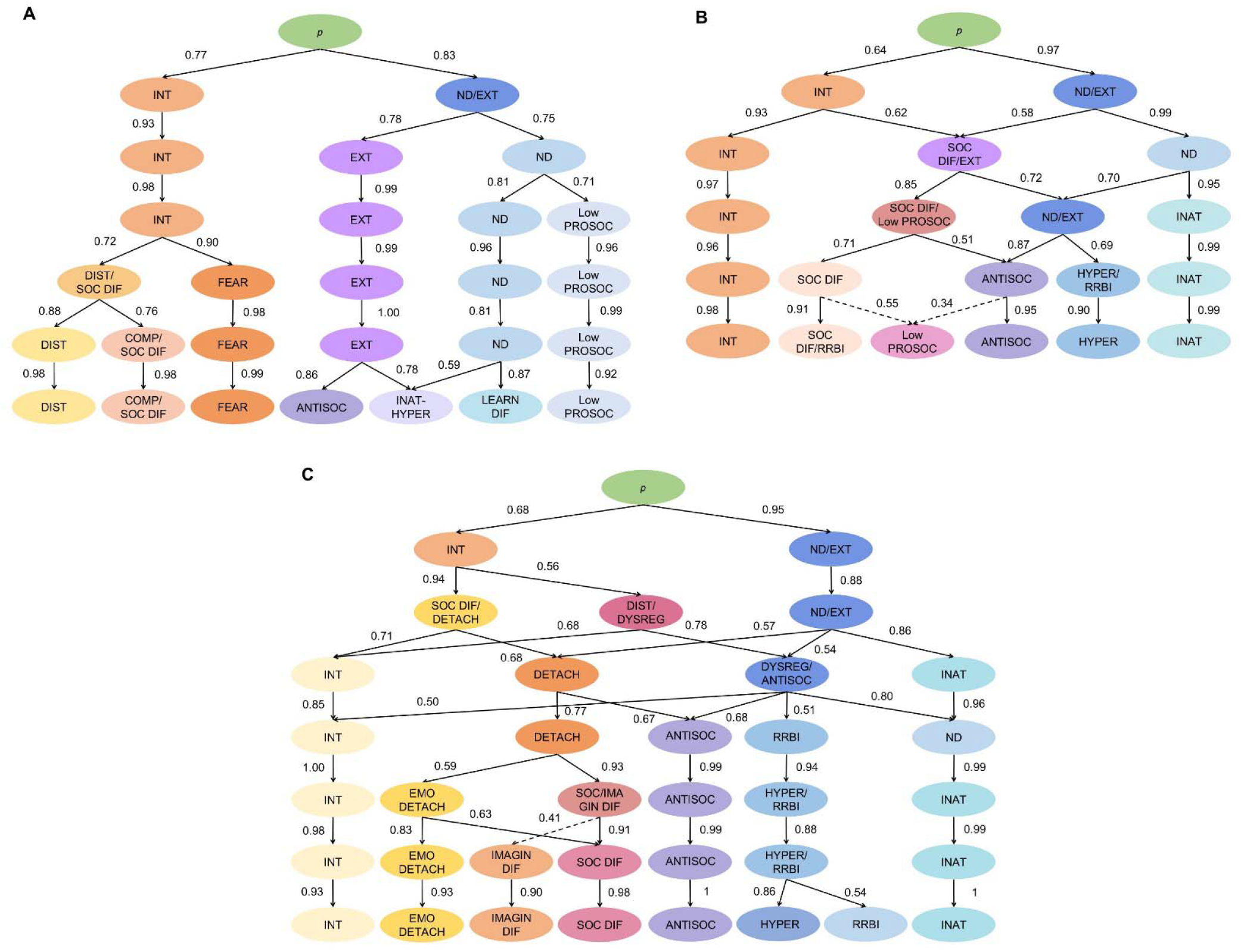
Phenotypic factor structure at age 7 (A), 12 (B), and 16 (C). Notes: Hierarchical structures are based on exploratory structural equation modelling (ESEM) of parent-reported neurodevelopmental traits and psychiatric symptoms. The displayed paths depict correlations of at least 0.50 with a shift of at least 1 primary loading (≥0.35) across levels, indicating movement of significant content from a higher level to a lower level of the hierarchy. The only exceptions were the dashed paths to the low prosociality factor in the 6-factor model at age 12 and to the imagination difficulty factor in the 7-factor model at age 16, where no paths from the higher-level model met these criteria, therefore the largest paths with a shift in at least 1 primary loading are shown. ANTISOC=antisociality, DETACH=detachment, DIST=distress, DYSREG=dysregulation, EMO DETACH=emotional detachment, EXT=externalizing, INAT=inattention, HYPER=hyperactivity-impulsivity, IMAGIN DIF=imagination difficulties, INT=internalizing, LEARN DIF=learning difficulties; Low PROSOC=low prosociality, ND=neurodevelopmental, p=general psychopathology, RRBI=repetitive and restrictive behaviors and interests, SOC DIF=social difficulties.

At age 7, the neurodevelopmental factor in the 3-factor solution primarily captured attention difficulties (e.g., poor attention span), low prosociality (e.g., low tendency to help or show empathy), and learning difficulties with writing, reading, coordination, and speech. This factor showed small correlations with externalizing (r=.14) and internalizing (r=.34) factors. The neurodevelopmental factor subsequently split into more specific inattention-hyperactivity, learning difficulties, and low prosociality factors in the final 7-factor solution. Other autistic traits (social difficulties, restricted and repetitive behaviors and interests [RRBIs]) clustered separately in a factor mainly emerging from a higher-order internalizing factor (Fig. 1A).

The age-12 neurodevelopmental factor in the 3-factor solution captured inattention, hyperactivity-impulsivity, and pragmatic language difficulties, and showed moderate-to-large correlations with internalizing (r=.44) and social difficulties/externalizing (r=.51) factors. In the final 6-factor solution, neurodevelopmental traits split across four factors encompassing inattention, hyperactivity-impulsivity, low prosociality, and social difficulties and RRBIs (Fig. 1B).

The age-16 neurodevelopmental factor in the 5-factor solution mainly encompassed attention difficulties (e.g., distractibility, poor attention switching, low perseverance), hyperactivity, and abstract thinking difficulties. Correlations with other factors in this solution ranged from negligible (r=-.01 with detachment) to moderate (r=.46 with antisociality). The final 8-factor solution showed the reorganization of neurodevelopmental traits into narrower factors, capturing inattention, hyperactivity-impulsivity, RRBIs, social difficulties, and imagination difficulties (Fig. 1C). Secondary analyses of self-reported items at age 16 revealed a neurodevelopmental factor in the 3-factor solution, including difficulties with attention, social interaction, motivation/perseverance, and hyperactivity, which split into distinct inattention, hyperactivity, and social difficulties factors in more differentiated solutions (Supplementary Results 4, Supplementary Fig. 1, Supplementary Table 8).

Overall, the neurodevelopmental factors across development captured a broad set of partly overlapping neurodevelopmental traits traditionally belonging to autism and ADHD. Despite differences in assessments (and hence, items) available across ages, especially at age 7 vs. 12/16, we found moderate-to-large associations between the neurodevelopmental factors over time (age 7-12 β=0.38, age 7-16 β=0.34, age 12-16 β=0.63; all p<0.001; Supplementary Table 9).

### Etiology of the neurodevelopmental spectrum (Q2)

Twin analyses of the extracted factor scores showed that the neurodevelopmental factor was highly heritable at each age (age-7 h^2^=0.60, age-12 h^2^=0.79, age-16 h^2^=0.82), with moderate-to-large overlap in genetic effects across development (age 7-12 r_g_=0.45, age 7-16 r_g_=0.41, age 12-16 r_g_=0.70). See Supplementary Results 5, Supplementary Tables 10-11, and Supplementary Fig. 2 for details.

Polygenic scores for neurodevelopmental disorders and cognitive and educational phenotypes (Supplementary Table 2) significantly predicted the neurodevelopmental factors, with significant effects explaining between 0.08-2.30% of variance (Fig. 2A, Supplementary Table 12). Effects of polygenic scores were particularly consistent across age-12 and age-16 neurodevelopmental factors based on parent-reports, mirroring the high phenotypic and genetic correlation between these factors. Some associations at age 7 were smaller or nonsignificant (e.g., polygenic scores for autism and childhood intelligence), whereas polygenic scores for Tourette syndrome were predictive at age 7 but not in later development, potentially reflecting the inclusion of motor difficulties captured by this factor in assessments at age 7 but not later waves. Exploratory multi-polygenic-score analyses showed that combining these 18 polygenic scores from primary analyses explained up to 3.36% of variance in the neurodevelopmental factors, with a non-significant increase in variance to up to 3.93% when combining a wider set of 35 polygenic scores also for other psychiatric and personality phenotypes (Supplementary Fig. 3, Supplementary Table 13).

**Fig. 2.**
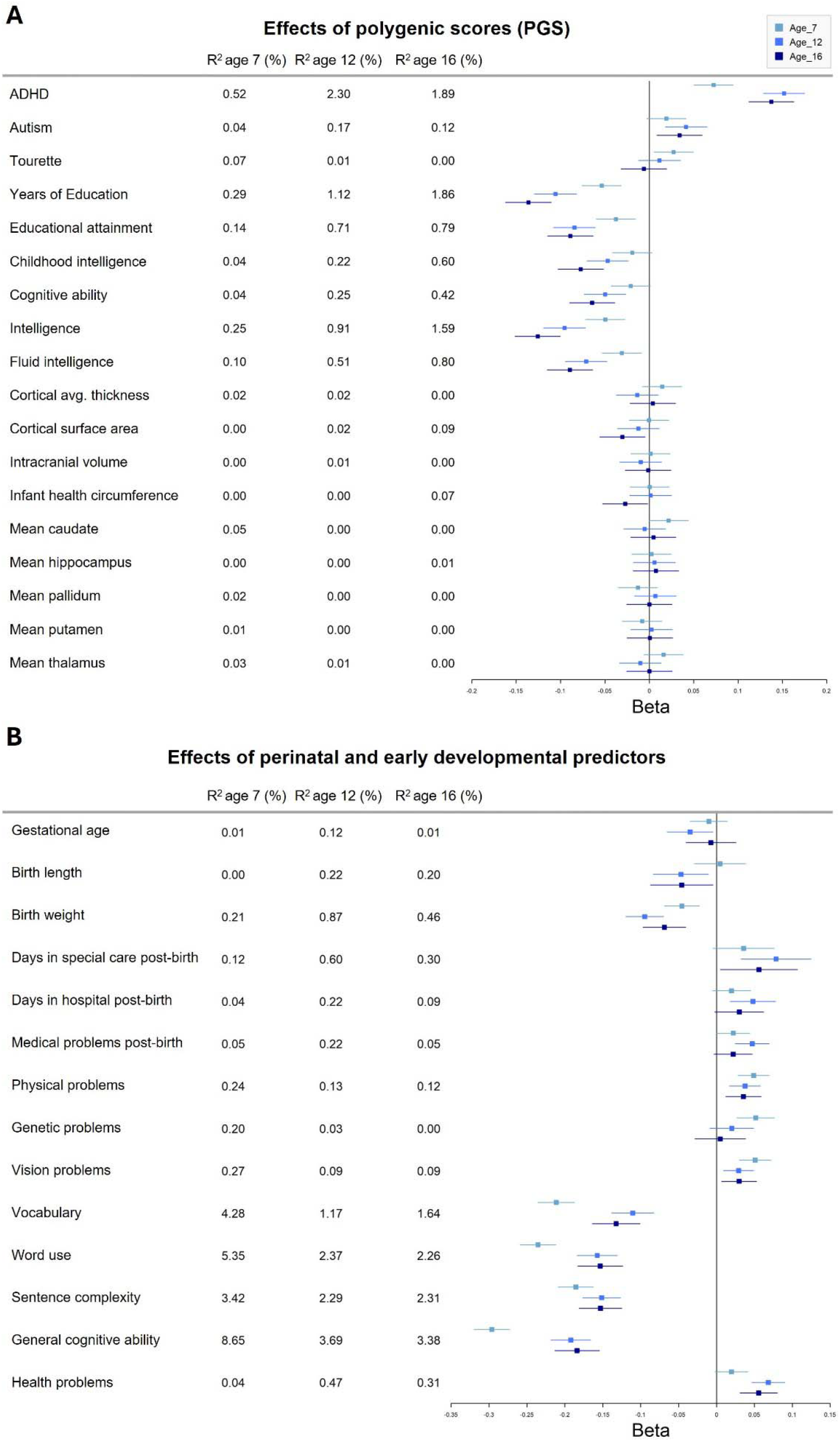
Effects of polygenic scores (A) and of perinatal/early developmental predictors (B) on neurodevelopmental factors at age 7, 12, and 16. Notes: Standard errors not crossing 0 indicate significant effects (p<.05). In panel A, polygenic scores were derived from available genome-wide association studies (GWAS) of neurodevelopmental conditions (ADHD, ASD, Tourette) and cognitive, educational, and brain phenotypes in independent samples. Further details about each measure can be found in Supplementary Table 2. In panel B, measures of vocabulary, word use, sentence complexity, general cognitive ability and health problems were collected at age 3 assessments, whereas other information was collected at first contact (around 18 months of age). Further details about each measure can be found in Supplementary Table 3.

Besides substantial genetic effects, twin analyses showed significant individual-specific environmental contributions at each wave (age-7 e^2^=0.40, age-12 e^2^=0.16, age-16 e^2^=0.18), with moderate developmental overlap (age 7-12 r_e_=0.36, age 7-16 r_e_=0.28, age 12-16 r_e_=0.40) (Supplementary Results 5, Supplementary Tables 10-11).

In analyses of specific perinatal/early developmental predictors, we found that neurodevelopmental factors were consistently predicted across time by lower scores on language acquisition (vocabulary, word use, sentence complexity) and cognitive ability at age 3 (R^2^=1.2-8.65%), as well as by low birth weight and physical and vision problems reported at 18 months (albeit with small effects, R^2^=0.05-0.27%). Most other predictors showed significant small effects at one or two waves (Figure 2B, Supplementary Table 14).

### Association with cognitive/educational outcomes (Q3)

The neurodevelopmental factor significantly predicted all cognitive and educational outcomes (i.e., general cognitive ability, academic achievement) across development, with both cross-sectional and longitudinal associations showing small-to-moderate beta coefficients (Fig. 3, Supplementary Table 15). Cross-sectionally, the neurodevelopmental factor explained 0.07-10.97% of variance in cognitive and educational outcomes at age 7, 5.18-9.37% at age 12, and 5.37-20.61% at age 16. Longitudinally, the age-7 factor explained 2.05-6.51% of the variance in age-12/16 outcomes, and the age-12 factor explained 3.43-10.06% of the variance in age-16 outcomes (Supplementary Table 15).

**Fig. 3.**
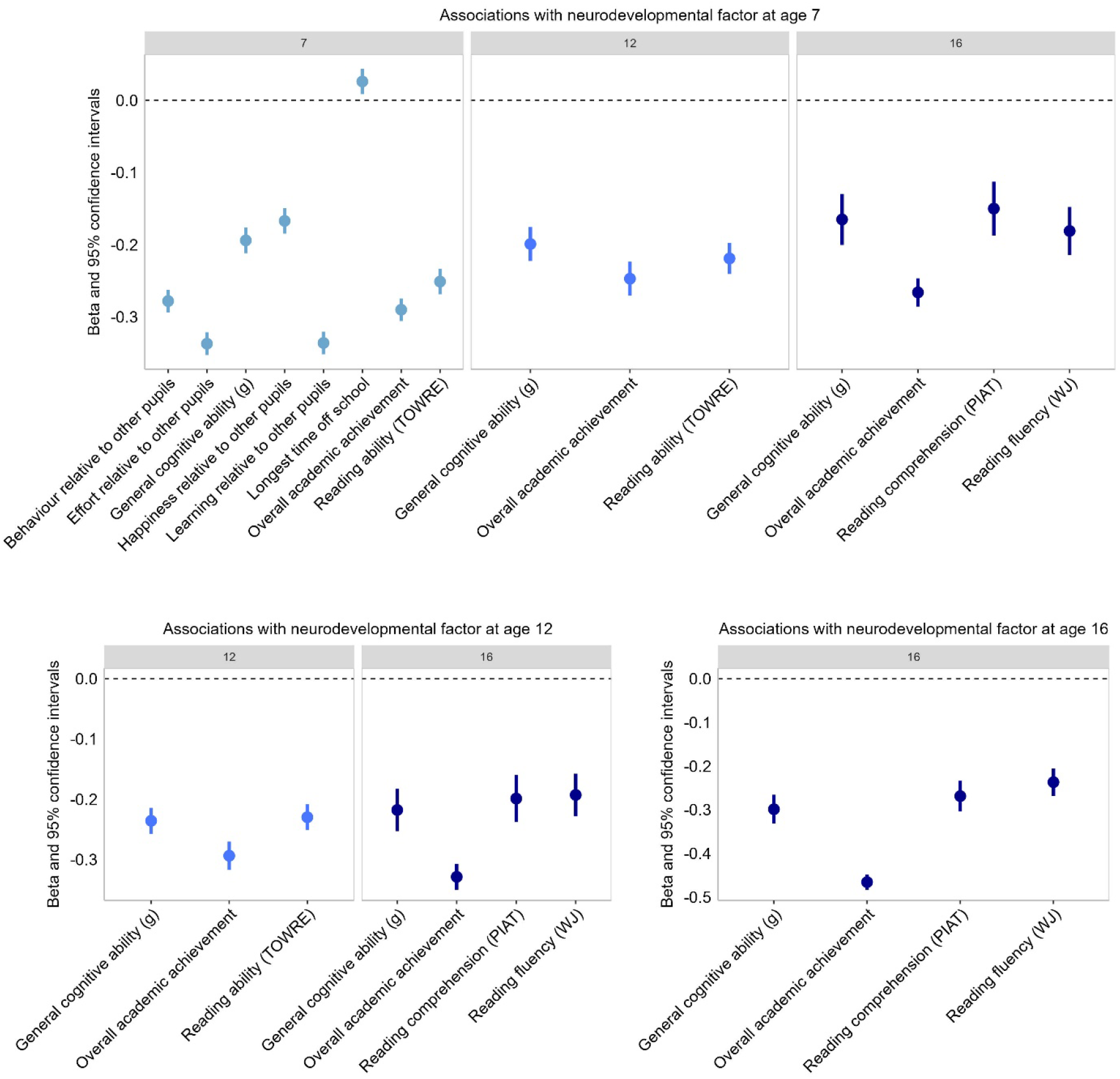
Effects of the neurodevelopmental factor on cognitive and educational outcomes, in cross-sectional and longitudinal analyses at age 7, 12, and 16. Notes: Effects of the age-7 factor on teacher-reported referral to special education support at age 7, and of the age-7 and age-12 factors on teacher-reported receipt of special education at age 12 are not displayed, as they showed large positive associations (Supplementary Table 15), unlike most of the other significant outcomes, which showed negative associations.

Bivariate twin models examining the genetic and environmental effects underlying these associations showed significant genetic overlap between the neurodevelopmental factor and all cognitive and educational outcomes (apart from a measure of longest time off school) (Supplementary Results 6, Supplementary Table 16). Positive genetic correlations between neurodevelopmental factors and special education support measures were substantial, ranging between 0.49-0.53, while negative genetic correlations, for example, between neurodevelopmental factors and general cognitive ability, ranged between -0.12 and -0.63.

### Specificity of associations (Q4)

Finally, we repeated random-intercept models addressing Q2 and Q3 controlling for other transdiagnostic factors in the 3-factor solution at ages 7 (internalizing and externalizing), 12 (internalizing and externalizing/social difficulties) and 16 for self-report data (internalizing and externalizing/psychosis), and in the 5-factor solution at age 16 for parent report data (internalizing, detachment, antisocial, and RRBIs). Most of the significant effects were slightly attenuated but remained significant (Supplementary Tables 12, 14, and 15 for details), suggesting that the neurodevelopmental factor showed specific associations with predictors and outcomes that were not explained by shared variance with other psychiatric dimensions.

## Discussion

We conducted the most comprehensive empirical investigation to date of the neurodevelopmental spectrum, focusing on its phenotypic structure, etiological validity, predictive utility, and specificity across development. Using rich data from a large nationally representative longitudinal study, we identified a neurodevelopmental spectrum consistently including features of ADHD and autism at ages 7, 12, and 16 years. This factor emerged alongside well-established transdiagnostic psychiatric spectra (e.g., internalizing and externalizing factors), extending current transdiagnostic and hierarchical dimensional models that have typically omitted neurodevelopmental traits [19, 20]. Leveraging complementary approaches to investigate etiological validity, we found that the neurodevelopmental spectrum was highly heritable across development and predicted by polygenic scores for neurodevelopmental conditions, cognition, and education, as well as by perinatal/early developmental predictors. Further, differences between children in the neurodevelopmental spectrum significantly predicted important functional outcomes (cognitive and educational performance), both concurrently and longitudinally, showing evidence of predictive utility. Finally, most of these associations with predictors and outcomes remained significant when accounting for better-established transdiagnostic spectra capturing co-occurring psychiatric symptoms, indicating that they were largely specific to the neurodevelopmental spectrum.

A key contribution of this study is identifying which features of neurodevelopmental conditions belong to the transdiagnostic neurodevelopmental spectrum across childhood and adolescence (Q1). Through hierarchical exploratory factor modeling of a comprehensive set of traits and symptoms collected at ages 7, 12, and 16, we demonstrated that this spectrum reflects the co-occurrence and shared features between neurodevelopmental conditions, extending previous literature that has mainly employed cross-sectional designs [7]. This spectrum emerged consistently as a distinct dimension, showing moderate correlations with other better-established transdiagnostic spectra, such as internalizing and externalizing spectra. In line with our hypotheses, the neurodevelopmental spectrum included features of all neurodevelopmental conditions investigated at each age. That is, we found that ADHD and autistic traits belong to this spectrum across all time points and across raters at age 16, together with learning and motor difficulties, which were only assessed in childhood. The neurodevelopmental spectrum consistently captured attention difficulties across development, which are a defining feature of ADHD but also associated with many other neurodevelopmental conditions (e.g., difficulties switching attention in autism) [13, 79]. This spectrum also encompassed socio-communication difficulties across childhood (e.g., low prosociality) and adolescence (e.g., pragmatic language difficulties), which are typical of autism but often displayed by individuals with other neurodevelopmental conditions [22, 80]. Interestingly, parent-reported hyperactivity and autistic RRBIs showed a high degree of clustering in adolescence, splitting into separate factors only in the most differentiated solutions. This suggests a transdiagnostic link between hyperactivity and RRBIs, potentially underpinned by shared genetic factors [9] or physiological arousal dysregulation [81, 82], as well as similarities between these behaviors that potentially make their distinction difficult using questionnaires. While our primary analyses were based on parent-reports, ADHD and autistic features showed a similar pattern in analyses of self-reports at age 16, as inattention, hyperactivity, and social difficulties also clustered in the self-reported neurodevelopmental spectrum. Taken together, our findings indicate that many neurodevelopmental traits across development can be conceptualized within a neurodevelopmental spectrum, supporting the value of collecting broad transdiagnostic assessments of neurodevelopmental traits in both research and clinical settings [7].

In addition to showing which traits fit within a neurodevelopmental spectrum, our findings provide new insights into the architecture of other transdiagnostic spectra. Hyperactivity-impulsivity traits of ADHD had a clear neurodevelopmental component across development, but were also partly captured by the externalizing spectrum (mainly including antisocial behavior, oppositionality, and callousness). This warns against considering ADHD a purely externalizing or neurodevelopmental condition, extending previous cross-sectional evidence [27, 40, 83]. Further, whereas many items assessing social difficulties were included in the neurodevelopmental spectrum, at least some autistic features (e.g., social interactions, RRBIs) showed partial contributions from internalizing dimensions, particularly in childhood. This pattern may be explained by difficulties disentangling autistic traits from social anxiety (e.g., “solitary or tends to play alone”) and obsessive-compulsive disorder (e.g., “fussy or over-particular”), which typically belong to internalizing dimensions, thereby driving high correlations between questionnaire items [84]. Finally, most positive and negative psychosis symptoms/traits (e.g., paranoia, blunted affect) at age 16, using either parent- or self-reports, were not included in the neurodevelopmental spectrum but rather in thought disorder and detachment dimensions, respectively. These novel findings support their current classification under these dimensions in the HiTOP framework, which is based on prior studies that did also neurodevelopmental traits [19, 41], but contrast with neurodevelopmental conceptualizations of psychosis [41, 42]. Together, these findings help us clarify where neurodevelopmental traits fit in transdiagnostic psychiatric frameworks and highlights the value of applying data-driven approaches to individual traits rather than heterogeneous diagnostic categories [6–8].

By focusing for the first time on both genetic and environmental influences through multiple complementary methodologies, our study sheds new light on the etiology and validity of the neurodevelopmental spectrum across development (Q2). High heritability in twin analyses and significant effects of polygenic scores converged in indicating that the neurodevelopmental spectrum has a strong genetic basis across childhood and adolescence, consistent with genetic studies of individual neurodevelopmental conditions and traits [3, 85]. These findings extend recent evidence that genetic liabilities for individual neurodevelopmental conditions are underpinned by a genomic neurodevelopmental factor [43–45]. Most of the polygenic scores showing significant effects explained a greater amount of variance in adolescence than in childhood, consistent with the higher twin heritability of the neurodevelopmental spectrum in adolescence than in childhood. A possible explanation is that this factor in adolescence mainly included ADHD and autistic traits, which typically show very high heritability estimates [3, 9, 85], whereas the childhood factor also included specific learning and communication difficulties, which are slightly less heritable [3]. This pattern might also be explained by potential contributions of non-additive genetic effects, not captured by polygenic scores, to the neurodevelopmental spectrum at age 7 but not 12-16, in line with our twin results. Future studies should test the possible role of rare genetic variants in the neurodevelopmental spectrum, extending work on individual neurodevelopmental diagnoses [86, 87]. Furthermore, perinatal and early developmental factors, such as low birth weight and language delays, also explained meaningful variance in the neurodevelopmental spectrum across development, although the potential role of familial/genetic confounds in these associations remains to be ruled out [51]. Secondary analyses at age 16 based on self-reports displayed broadly consistent findings, despite showing lower heritability and higher individual-specific environmental effects than the parent-report factor, consistent with twin studies of self-reported ADHD and autistic traits [10, 88]. Since individual-specific effects include measurement error in twin modelling, these findings suggest that parent-reports may be more valid and reliable than self-reports for delineating the neurodevelopmental spectrum in adolescence. These findings confirm the hypothesized contribution of genetic predispositions and early-life exposures to the neurodevelopmental spectrum [7] and support its validity for research and clinical practices.

Our study further provides novel evidence for the predictive utility of the neurodevelopmental spectrum (Q3). We found that the spectrum explains up to 21% of variance in real-world functional outcomes, both concurrently and longitudinally. Children and young people with higher neurodevelopmental spectrum scores showed worse outcomes, particularly lower educational performance and greater support needs throughout primary and secondary education relative to their peers. These findings contribute to an emerging body of evidence from longitudinal studies showing substantial predictive utility of psychiatric dimensions, including dimensional measures of neurodevelopmental traits [47, 48]. Through twin analyses, we also showed that these associations were underpinned by significant genetic effects, consistent with previous studies on the association between individual neurodevelopmental diagnoses and cognitive/educational outcomes [55, 56, 89]. The strong predictive power of the neurodevelopmental spectrum suggests that assessing broad neurodevelopmental traits early in development could be particularly useful for identifying children at risk for poor functional trajectories, who would benefit from early support, particularly in school settings.

Besides identifying associations with important predictors and outcomes, we finally examined whether these associations are specific to the neurodevelopmental spectrum or shared with other transdiagnostic dimensions (Q4). We found that the reported associations remained robust, albeit slightly reduced, when controlling for other transdiagnostic dimensions capturing internalizing, externalizing, and psychosis symptoms. This suggests that the neurodevelopmental spectrum is largely independent of other transdiagnostic dimensions and shows specific associations with key predictors and outcomes. Together, our findings indicate that the neurodevelopmental spectrum is a coherent, valid, and useful dimension that consistently emerges alongside well-established transdiagnostic dimensions, uniquely accounts for neurodevelopmental traits, and predicts important functional outcomes over and above other transdiagnostic dimensions.

These findings have important implications for both research and clinical practice. By providing novel evidence on the neurodevelopmental spectrum, our work advances contemporary psychiatric classification and provides empirical support for revising transdiagnostic psychiatric frameworks, such as HiTOP [19], to incorporate a neurodevelopmental spectrum alongside established transdiagnostic dimensions. Integrating neurodevelopmental traits within such frameworks through this spectrum yields novel dimensional phenotypes for investigating underlying mechanisms and developing biomarkers, moving beyond current approaches centering individual heterogeneous diagnoses [35–38]. Critically, this perspective further offers a novel conceptual framework for recognizing the common overlap and continuity between neurodevelopmental and other psychiatric traits across the life course [7]. Our findings highlight that clinicians should assess neurodevelopmental traits as well as symptoms of other psychiatric conditions, with the potential to reduce diagnostic overshadowing and delayed support. Finally, our work indicates the need to develop new transdiagnostic, more holistic assessment approaches in clinical settings, as well as intervention and support strategies that address broad neurodevelopmental difficulties and prevent adverse developmental trajectories [7, 90].

This study has the following limitations. First, while the TEDS participants included in this study are reasonably representative of the UK population at the time of recruitment, they are less representative of the current UK population, which is more diverse. For example, TEDS predominantly includes individuals who identify as White, consistent with the UK population in the mid-1990s [50]. Future research should extend this work to more diverse samples, in line with calls for increasing diversity in transdiagnostic and genetic research [91, 92]. Second, while our study assessed the largest number of neurodevelopmental traits longitudinally to date among studies investigating their structure and placement in transdiagnostic models, it provided limited coverage beyond ADHD and autistic features in adolescence. Additionally, some of the neurodevelopmental traits included at age 7 were based on items developed by the TEDS team to assess parent-reported diagnoses or difficulties based on DSM-IV criteria. Future studies should extend this work by assessing all traits of neurodevelopmental conditions alongside other psychiatric symptoms across development, and using validated assessments based on current DSM-5 criteria. Third, due to differences in the measures collected across waves, factor analyses included only partly overlapping neurodevelopmental traits at ages 7, 12, and 16, meaning that we could not clarify whether differences between factors across waves arose from differences in measures or genuine developmental effects. Similarly, differences in assessments across parent- and self-reports did not allow formal examination of rater effects. Nevertheless, we identified clear similarities in findings across time and raters, showing robust effects despite assessment differences. Fourth, polygenic scores represent a proxy for individual genetic predisposition, but only capture common genetic variants and their predictive power depends on the power of the genome-wide association studies (GWAS) they were derived from. This may explain their generally modest associations with the neurodevelopmental spectrum and nonsignificant effects of polygenic scores of brain phenotypes, based on smaller (and hence less powered) GWAS relative to GWAS for psychiatric and cognitive/educational phenotypes [62, 63]. As larger GWAS are conducted, future studies will further clarify the genetic underpinnings of the neurodevelopmental spectrum. Finally, we used a single dataset due to a lack of comparable samples, and the majority of analyses did not employ a cross-validation approach, meaning that future research should replicate our findings.

To conclude, by using a well-established, comprehensively phenotyped, genetically sensitive longitudinal sample, our study contributes novel and robust evidence on the neurodevelopmental spectrum across development. Our findings highlight the relevance of this transdiagnostic spectrum for understanding the complexity of neurodevelopmental conditions and their functional outcomes. Building on these findings, future efforts should develop broader transdiagnostic assessments and formally integrate the neurodevelopmental spectrum into transdiagnostic dimensional classifications to fully realize their potential for advancing research and clinical care. This will encourage joint recognition of neurodevelopmental traits and co-occurring and later-emerging psychiatric symptoms, with an eye to more personalized and holistic intervention strategies.

## Supporting information

SupplMethods&Results

## Acknowledgements

The authors would like to express their gratitude to the participants and families taking part in the Twins Early Development Study (TEDS). The authors gratefully acknowledge the dedicated TEDS team for their commitment to the project. TEDS is funded by a program grant from the Medical Research Council (MR/V012878/1 to Professor Thalia C. Eley and previously MR/M021475/1 to Professor Robert Plomin), with additional support from Medical Research Council grant G1100559 (to Professor Angelica Ronald). Dr Michelini was partly funded by a Klingenstein Third Generation Foundation fellowship (20212999). Dr Malanchini was supported by a Jacobs Foundation Research Fellowship. Drs Malanchini and Michelini were further supported by Medical Research Council grant UKRI1506. Wangjingyi Liao and Shiqi Lu were supported by Chinese Scholarship Council PhD studentships. Chiara Caserini was supported by an Economic and Social Research Council London Interdisciplinary Social Sciences Doctoral Training Partnership PhD studentship. Dr Rimfeld was supported by a Sir Henry Wellcome Postdoctoral Fellowship by the Wellcome Trust (213514/Z/18/Z) and Medical Research Council grant (UKRI1503). For the purpose of open access, the author has applied a CC BY public copyright licence to any Author Accepted Manuscript version arising from this submission.

## Author Contributions

Conceptualization: Michelini

Data acquisition: Eley, Ronald

Formal analysis: Michelini, Rimfeld, Liao, Lu

Supervision: Michelini, Rimfeld, Malanchini

Visualization: Michelini, Caserini, Liao, Malanchini

Interpretation: All authors

Writing - Original Draft: Michelini

Writing - Review & Editing: All authors

## Data availability

Researchers can apply for access to the TEDS data through their data access mechanism (www.teds.ac.uk/researchers/teds-data-access-policy).

## Code availability

Code is available on Open Science Framework alongside the study preregistration (https://osf.io/dqbnj/?view_only=f2510b6766bc4b86bf9344fee1139233).

