## Supplementary material for "The neurodevelopmental spectrum: phenotypic architecture, etiology, predictive utility, and specificity across development": SupplMethods&Results

**SUPPLEMENTARY METHODS**

**1. Deviations from the pre-registration**

Our research aims, hypotheses, and analysis plan were pre‐registered on the Open Science Framework before accessing the data (<https://osf.io/dqbnj/?view_only=f2510b6766bc4b86bf9344fee1139233>). We made small deviations from our pre-registered analysis plan related to data availability and for completeness. First, while the pre-registration states that analyses would be conducted on measures collected across raters, parent ratings were used in the main analyses because parents completed the largest number of relevant assessments across all three waves, whereas self- and teacher-reports were available only for a subset of measures. Separate additional analyses were conducted on the age 16 self-report ratings. Second, analyses on age 16 parent-report data included the abbreviated Autism Spectrum Quotient (AQ) scale and the Eating Disorder Diagnostic Scale (EDDS), which were accidentally omitted from the preregistration. Third, the analyses of age 16 self-report data did not include items about alcohol, tobacco, cannabis and drug use and from the Hypomania checklist (HCL-16) scales (included in the pre-registration) because these data were collected only in a minority of participants completing other assessments at this wave. Fourth, analyses on the temporal stability of the neurodevelopmental spectrum also included phenotypic mixed models looking at associations between neurodevelopmental factors over time, controlling for twin relatedness, age, and sex, in addition to equivalent twin analyses mentioned in the preregistration. Fifth, analyses looking at associations with cognitive and educational outcomes also included twin modelling to estimate genetic and environmental effects on these associations, mirroring the phenotypic analyses. Sixth, in analyses of genetic and early environmental/developmental predictors, we did not include the cross-disorder ADHD-ASD-TS-OCD polygenic score because summary statistics were not publicly available, nor the TEDS perinatal outliers’ composites as these outliers reflect binary variables derived from continuous variables included in our analyses, and were therefore redundant.

**2. DNA collection, genotyping, and polygenic scores**

DNA samples have been obtained from 12,500 individuals and genotyped on one of the two DNA microarrays (Affymetrix GeneChip 6.0 or Illumina HumanOmniExpressExome chips). After stringent quality control, the total number of TEDS participants available for genomic analyses was 10,346 (including 7,026 unrelated individuals and 3,320 additional dizygotic [DZ] co-twins). Of these, 7,289 individuals were genotyped on Illumina arrays, and 3,057 individuals were genotyped on Affymetrix arrays (see [1] for a detailed description). Genomic data were available for a subsample of participants included at age 7 (N=8,262), 12 (N=7,080), and 16 (N=6,030).

We calculated the polygenic scores using LDpred and, since 2022, LDpred2-auto [2, 3] based on the largest available genome-wide association study (GWAS) for each phenotype of interest (Supplementary Table 2). All available SNPs were included in the polygenic scores (i.e., p-value threshold of 1), in line with previous work in this sample [4–6]. Further information on individual GWAS and summary statistics can be found elsewhere (<https://www.med.unc.edu/pgc/download-results/>; <https://www.teds.ac.uk/datadictionary/studies/measures/polygenic_scores.htm#list>)

**3. Twin analyses**

The twin method offers a natural experiment to estimate the proportion of phenotypic variance of a trait or covariation between traits explained by genetic and environmental effects, using the known genetic relatedness between monozygotic (MZ) and dizygotic (DZ) twin pairs. MZ twins share 100% of their segregating genes, while DZ twins share on average 50% of their segregating genes. Both MZ and DZ twin pairs share 100% of the environmental effects of growing up in the same family. Twin model-fitting analyses can decompose phenotypic variance into contributions of additive genetic effects (A), common environmental effects (C) which make members of a twin pair similar to one another, and individual-specific environmental effects (E) which make members of a twin pair differ, including measurement error. If MZ correlations on trait similarity are larger than DZ correlations, then genetic influences on a trait can be assumed. In the event of twin correlations in MZ twins more than twice as large as those in DZ twins, indicating non-additive genetic effects, models decomposing A, non-additive genetic effects (D) and E effects are tested instead, as C and D cannot be simultaneously estimated [7]. Twin analyses were run in the R package OpenMx [8] using the standardized residuals after accounting for the effects of sex and age, as is standard practice in quantitative genetic model-fitting [9].

**4. Single-polygenic-score vs. multi-polygenic-score analysis**

A pre-registered exploratory analysis compared a single-polygenic-score with a multi-polygenic-score approach for predicting the neurodevelopmental factor across development [4, 5, 10]. Specifically, we compared the predictive power (i.e., variance explained) in the neurodevelopmental factor explained by the most predictive polygenic score at each age (ADHD polygenic score) (model 1), jointly by the 18 polygenic scores included in our primary analyses (model 2), and jointly by these 18 polygenic scores and 17 additional polygenic scores for other psychiatric disorders and associated traits (e.g., personality) (Supplementary Table 2). These analyses were run using elastic net models, which are designed to model linear relationships with the penalties of both LASSO and ridge regression, allowing us to manage multicollinearity and enhance model prediction accuracy. We ran elastic net models using the *caret* and *glmnet* packages in R, optimizing the hyper-parameters through tuning and 10-fold cross-validation. For these analyses, unlike other analyses using mixed models to account for family relatedness, we randomly selected one twin per family to avoid possible overfitting due to the inclusion of related individuals. To compare the predictive power of the most predictive single polygenic score (ADHD, model 1) and models including multiple polygenic scores (models 2 and 3), we used bootstrapping with 1000 iterations for each analysis. This involved generating multiple resamples of the data to estimate the distribution of the R-squared values for both the single PGS and the elastic net models. The comparison aimed to determine whether the multi-polygenic-score approach leveraging multiple predictors simultaneously offers a significant advantage over single polygenic score predictions in explaining the variance in the neurodevelopmental factors. Non-overlapping CI indicated a significant increase in predictive power (Supplementary Figure 3).

**SUPPLEMENTARY RESULTS**

**1. Hierarchical dimensional structure in childhood (age 7, parent-report)**

Prior to analyses, we checked for items with very low endorsement (>99.5% of participants had 0 responses) and with very high correlations (r>.85), and removed diagnosed autism due to its low endorsement causing model non-convergence, following standard analytic procedures applied in previous work [11–13]. A total of 74 items from age 7 assessments were included in ESEM and parallel analyses. Parallel analyses indicated that up to 15 factors could be extracted. As parallel analysis tends to over-factor, we also examined the interpretability of these factor solutions, defined as the presence of at least 4 clear primary loadings (highest loading ≥0.35 and at least 0.10 greater than all other loadings) for each factor, in line with previous work [13–15]. Solutions including 1 to 7 factors were acceptable (Supplementary Table 5). Solutions with more than 7 factors showed poor interpretability as each included at least one factor with only three or fewer primary loadings.

Using Goldberg’s bass-ackwards approach[16], models including 1 to 7 factors were arranged in a hierarchical structure, with paths linking across levels representing large associations (Figure 1). The 1-factor structure reflected a general psychopathology factor, or p, with strong loadings on most items (Supplementary Table 5). The 2-factor solution showed broad internalizing and neurodevelopmental/externalizing factors. In the 3-factor solution, the neurodevelopmental/externalizing factor split into separate factors capturing neurodevelopmental traits (including attention, learning and motor difficulties, and low prosociality) and externalizing behaviors (including hyperactivity-impulsivity and antisocial behavior). In the 4-factor structure, a factor capturing low prosociality (e.g. inconsiderate, unhelpful, unking to other children) emerged from the neurodevelopmental factor. In the 5-factor solution, the broad internalizing factor split into distress/social difficulties (e.g., solitary, tendency to blame themselves, and be self-critical) and fear (e.g., nervous/clingy, afraid of social situations, shy/timid) factors. In the 6-factor structure, the distress/social difficulties factor split into a more defined distress factor (e.g., worries, tendency to blame themselves, self-critical) and a factor capturing social difficulties and repetitive and restricted behaviors and interests (e.g., different social interactions, unusual interests). Finally, in the 7-factor structure, an inattention-hyperactivity factor (capturing ADHD symptoms) emerged from content migrating from the externalizing factor and the neurodevelopmental factor; the remaining externalizing content formed a narrower antisocial behavior factor (e.g., tendency to lie/cheat, and consider themselves more important than others), whereas the remaining neurodevelopmental content delineated a factor capturing learning difficulties (e.g., difficulties with writing, reading, math, coordination, speech).

**2. Hierarchical dimensional structure in early adolescence (age 12, parent-report)**

No items showed very low endorsement (>99.5% of participants had 0 responses) or very high correlations (r>.85), thus a total of 104 items from age 12 assessments were included in ESEM and parallel analyses. Parallel analyses indicated that up to 20 factors could be extracted. Solutions including 1 to 6 factors were acceptable based on interpretability (Supplementary Table 6). Solutions with more than 6 factors showed poor interpretability as each included at least one factor with only three or fewer primary loadings.

Using Goldberg’s bass-ackwards approach [16], models including 1 to 6 factors were arranged in a hierarchical structure, with paths linking across levels representing large associations (Figure 1). A p factor at the apex in the 1-factor structure split into internalizing and neurodevelopmental/externalizing factors in the 2-factor solution. In the 3-factor solution, the neurodevelopmental/externalizing factor split into separate neurodevelopmental (e.g., inattention, hyperactivity-impulsivity, lack of planning, pragmatic language difficulties) and a broad factor encompassing externalizing behavior and social difficulties (e.g., unhelpful if someone is hurt, lies or cheats, does not chat spontaneously). In the 4-factor structure, the latter factor split into a factor encompassing social difficulties and low prosociality (e.g., unhelpful if someone is hurt, inconsiderate, does not chat spontaneously) and a neurodevelopmental/externalizing factor with additional contributions from the neurodevelopmental factor from the 3-factor model (e.g., very one sided, interrupts or intrudes, lies or cheats), whereas the remaining content in the neurodevelopmental factor formed an inattention factor. In the 5-factor solution, a more delineated social difficulties factor (broadly capturing social interaction and communication problems) emerged, and the neurodevelopmental/externalizing factor split into a factor capturing hyperactivity-impulsivity and restrictive and repetitive behaviors and interests (RRB) (e.g., motor overactivity, attention/memory for details) and an antisocial behavior factor, with the latter also including low prosociality from the social difficulties/low prosociality factor from the 4-factor structure. Finally, the 6-factor structure saw the emergence of a new factor capturing low prosociality (e.g., unhelpful, unkind) from the reorganization of items cross-loading onto the social difficulties and antisocial factors in the 5-factor structure, alongside other factors already included in the 5-factor structure. Note that repetitive and restricted interests and behaviors clustered with social difficulties in the 6-factor solution, rather than with hyperactivity-impulsivity.

**3. Hierarchical dimensional structure in mid-adolescence (age 16, parent-report)**

No items showed very low endorsement (>99.5% of participants had 0 responses) or very high correlations (r>.85), thus a total of 130 items from age 16 parent-report assessments were included in ESEM and parallel analyses. Parallel analyses indicated that up to 19 factors could be extracted. Solutions including 1 to 8 factors were acceptable based on interpretability (Supplementary Table 7). Solutions with more than 8 factors showed poor interpretability as each included at least one factor with only three or fewer primary loadings.

Using Goldberg’s bass-ackwards approach [16], models including 1 to 8 factors were arranged in a hierarchical structure, with paths linking across levels representing large associations (Figure 1). Similar to the hierarchical structures at ages 7 and 12, the 1-factor structure (p factor) split into internalizing and neurodevelopmental/externalizing factors in the 2-factor structure. In the 3-factor structure, the broad internalizing factor split into a social difficulties/detachment factor (including shyness, social withdrawal, apathy and affective flattening) and a distress/dysregulation factor (including depression symptoms and motor/emotional dysregulation). In the 4-factor structure, the neurodevelopmental/externalizing factor split into an inattention factor (e.g., lack of perseverance, difficulties planning, distractibility), a factor capturing dysregulation and antisocial behavior (e.g., motor overactivity, emotional dysregulation, fighting, bullying), and a factor capturing features of detachment (e.g., social withdrawal, apathy and affective flattening). The dysregulation/antisocial factor was strongly associated with the higher-level distress/dysregulation factor, and the detachment factor with the higher-level internalizing/detachment factor. In the 5-factor structure, the neurodevelopmental/externalizing factor split into a neurodevelopmental factor including attention difficulties, hyperactivity, lack of perseverance, and difficulties with creative thinking/imagination, an RRBI factor (e.g., fascinated by dates/numbers/patterns), and an antisocial behavior factor also including contributions from the detachment factor (e.g., callous, inconsiderate, tendency to lie/cheat), with the remaining content migrating in the internalizing factor. In the 6-factor structure, the detachment factor split into factors capturing social and imagination difficulties (e.g., difficulties with social interactions and mentalizing) and emotional detachment (e.g., affective flattening). In the 7-factor structure, the social/imagination difficulties factor formed a more delineated social difficulties factor merging with the content of the emotional detachment factor, with the remaining content in the social difficulties factor forming a factor capturing imagination difficulties (e.g., with creative thinking and mentalizing). Finally, the 8-factor structure saw the delineation of two separate factors capturing hyperactivity-impulsivity and RRBI from the factor including these behaviors in the 7-factor structure.

**4. Hierarchical dimensional structure in mid-adolescence (age 16, self-report)**

No items showed very low endorsement (>95% of participants had 0 responses) or very high correlations (r>.85), thus a total of 180 items from age 16 self-report assessments were included in ESEM and parallel analyses. Parallel analyses indicated that up to 17 factors could be extracted. Solutions including 1 to 14 factors were acceptable based on interpretability (Supplementary Table 8). Solutions with more than 14 factors showed poor interpretability as each included at least one factor with only three or fewer primary loadings.

Using Goldberg’s bass-ackwards approach [16], models including 1 to 14 factors were arranged in a hierarchical structure, with paths linking across levels representing large associations (Supplementary Figure 1). The general factor (p factor) included in the 1-factor solution split into an internalizing/thought factor, primarily capturing internalizing problems (e.g., anxiety and depression) and psychotic symptoms (e.g., paranoid ideation, hallucinations), and a broad neurodevelopmental factor, capturing attention difficulties, hyperactivity, social difficulties, anhedonia, and low prosociality in the 2-factor solution. In the 3-factor solution, the internalizing/thought factor split into a more defined internalizing factor, primarily capturing anxiety and depression problems, and a thought/externalizing factor, capturing psychotic symptoms as well as antisocial behaviors. In the 4-factor structure, the psychotic symptoms in the thought/externalizing factor migrated into a more defined thought factor, encompassing paranoid ideation, grandiosity, hallucinations, and RRBI (e.g., fascination for numbers/patterns). The remaining externalizing content in the thought/externalizing factor merged with inattentive/hyperactive features from the neurodevelopmental factor from the 3-factor structure into an inattention/dysregulation factor, capturing inattention and behavioral dysregulation (e.g., hyperactivity-impulsivity, antisocial behavior). The remaining content in the neurodevelopmental factor from the 3-factor solution formed a detachment factor in the 4-factor solution, capturing features of anhedonia, apathy, emotional detachment, social withdrawal, and difficulties in social occasions. This detachment factor also included contributions from the internalizing factor from the 3-factor structure. In the 5-factor structure, the thought factor split into paranoia factor and a grandiosity/RRBI factor. The detachment factor generated a narrower detachment factor, with remaining content in this factor merging with the internalizing factor. In the 6-factor structure, the grandiosity/RRBI factor from the 5-factor structure split into a more delineated grandiosity/antisocial factor, also capturing features of antisocial behavior, and an RRBI factor. In the 7-factor structure, the detachment factor from the 6-factor structure split into an anhedonia factor, capturing lack of pleasure and apathy, and a social difficulties factor, capturing social withdrawal and difficulties in social occasions. The RRBI factor in this structure also included hallucinations, and was thus labelled RRBI/hallucinations factor. In the 8-factor structure, the internalizing factor split into a distress/eating factor, primarily capturing depression and eating problems, and a fear factor, capturing anxiety sensitivity and phobias. Features of lack of motivation/effort included in the inattention/dysregulation factor from the 7-factor solution merged with the anhedonia factor, whereas the remaining content in the inattention/dysregulation factor formed a more delineated inattention-hyperactivity factor. In the 9-factor solution, the RRBI/hallucination factor from the 8-factor solution split into separate RRBI and hallucinations factors. Features of low prosociality moved from the social difficulties factor to a factor capturing anhedonia and low prosociality. Eating problems migrated from the distress/eating factor into a fear/eating factor. In the 10-factor structure, a new emotional detachment factor emerged from the reorganization of content from the RRBI and anhedonia/low prosociality factors in the 9-factor solution. Content from the latter factor further merged with content from the grandiosity factor into a grandiosity/antisociality factor. In the 11-factor structure, the latter factor split into more delineated grandiosity and antisociality factors. In the 12-factor structure, the inattention-hyperactivity factor split into separate inattention and hyperactivity-impulsivity factors. In the 13-factor structure, an eating factor emerged alongside distress and fear factors. Finally, in the 14-factor structure, the antisociality factor split into a psychopathy factor, capturing callousness and aggression, and a low prosociality factor, capturing a low tendency to be helpful and kind.

**5. Genetic and environmental effects on the neurodevelopmental factor**

Univariate Cholesky analyses on the neurodevelopmental factor at each time point (age 7, 12, and 16 years) using parent-reports. Results are presented in Supplementary Table 10. Twin correlations suggested the presence of non-additive (D) genetic effects on the neurodevelopmental factor at age 7 but not at age 12 and 16. We therefore fitted both ACE and ADE models at each age for completeness and to aid comparability of results. Both ACE and ADE models yielded similar results at each age, as we found similar estimates of broad heritability when comparing A effects in ACE models (age 7=0.599, age 12=0.791, age 16=0.824) with the sum of A+D effects (age 7=0.637, age 12=0.841, age 16=0.824) in ADE models. The rest of the variance was explained by modest individual-specific environmental effects. Secondary analyses on the neurodevelopmental factor at age 16 based on self-reports revealed that additive genetic (A=0.516) and individual-specific environmental (E=0.484) effects explained roughly equivalent proportions of variance in this factor (Supplementary Table 10).

Multivariate twin modelling was used to examine the overlap in genetic and environmental effects on the parent-reported neurodevelopmental factors across time (Supplementary Table 11). Cholesky models including A and E factors (AE models) were specified instead of ACE or ADE models, since we found negligible C effects across ages and there was an indication of possible D effects only at age 7 data. Results are provided in Supplementary Table 10 and Supplementary Figure 2. We found a moderate to large degree of overlap in genetic factors across time points. Genetic correlation between age 7 and age 12 factors was r_g_=0.450, between age 7 and 16 was r_g_=0.408, and between age 12 and 16 was r_g_=0.703. From these models we also found that 20.048% (i.e., 0.167/(0.167+0.666)) and 16.585% (i.e., 0.135/(0.135+0.277+0.402)) of the genetic effects on the neurodevelopmental factors at age 12 and 16, respectively, were shared with those on the age 7 factor, and that 40.795% (i.e., 0.277/(0.277+0.402)) of the heritability at age 16 with those at age 12. Estimates of individual-specific environmental correlations and shared individual-specific environmental effects across time were generally smaller, suggesting that environmental effects were largely time-specific. Secondary AE bivariate analyses also examined the overlap in genetic and environmental effects on the parent-reported and self-reported neurodevelopmental factors at age 16. Results indicated modest genetic (r_g_=0.258) and individual-specific environmental (r_e_=0.103) correlations (Supplementary Table 11).

**6. Genetic and environmental effects on the covariation of the neurodevelopmental factors with cognitive/educational outcomes**

Bivariate twin models were run to examine the cross-sectional and longitudinal covariation between the neurodevelopmental factor and cognitive and educational outcomes, mirroring phenotypic analyses. Analyses involving binary outcomes (referral for special education at age 7 and statement of special education needs at age 12) were carried out using a liability threshold model framework, which assumes that the liability of the binary measures is underpinned by a normally distributed continuum of risk [7, 17]. For most outcomes, we found evidence of significant genetic and individual-specific environmental correlations between the neurodevelopmental factor and cognitive/educational outcomes (Supplementary Table 16).

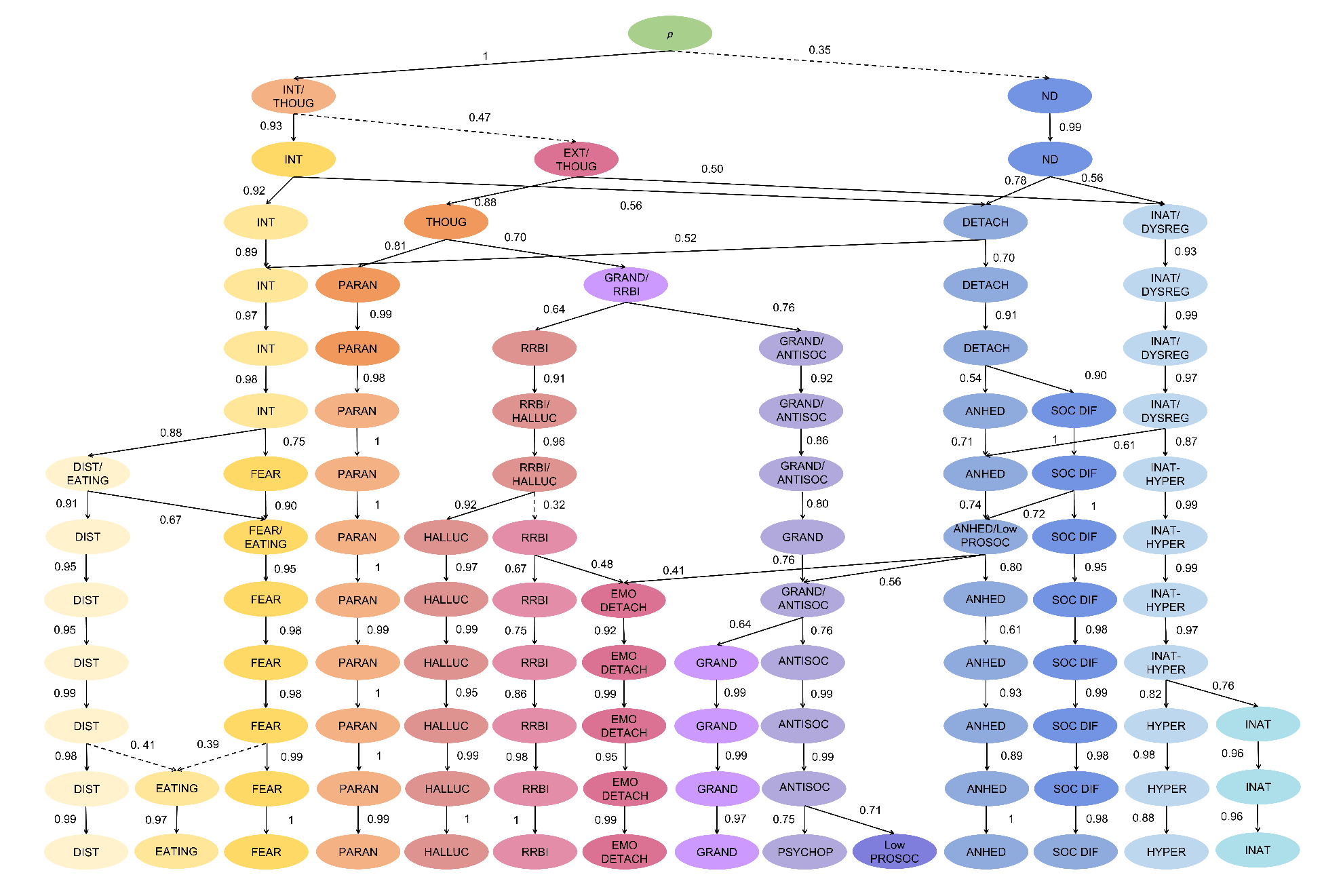
**Supplementary Figure 1**. Phenotypic factor structure at age 16 using self-reported items. Paths depict correlations of at least .50 with a shift of at least 1 primary loading across levels (indicating movement of significant content from a higher level to a lower level of the hierarchy); with the exception of the dashed paths where no paths met these criteria, therefore the largest path with a shift in at least 1 primary loading is shown. ANHED=anhedonia, ANTISOC=antisociality, DETACH=detachment, DIST=distress; DYSREG=dysregulation, EMO DETACH, emotional detachment, EXT=externalizing, GRAND=grandiosity, HALLUC, hallucinations, INAT= inattention, HYPER=hyperactivity, IMAGIN DIF=imagination difficulties, INT=internalizing, Low PROSOC=low prosociality, ND=neurodevelopmental, p=general psychopathology, PARAN=paranoia, PSYCHOP=psychopathy, RRBI =repetitive and restrictive behaviors and interests, SOC DIF=social difficulties, THOUG=thought disorder.

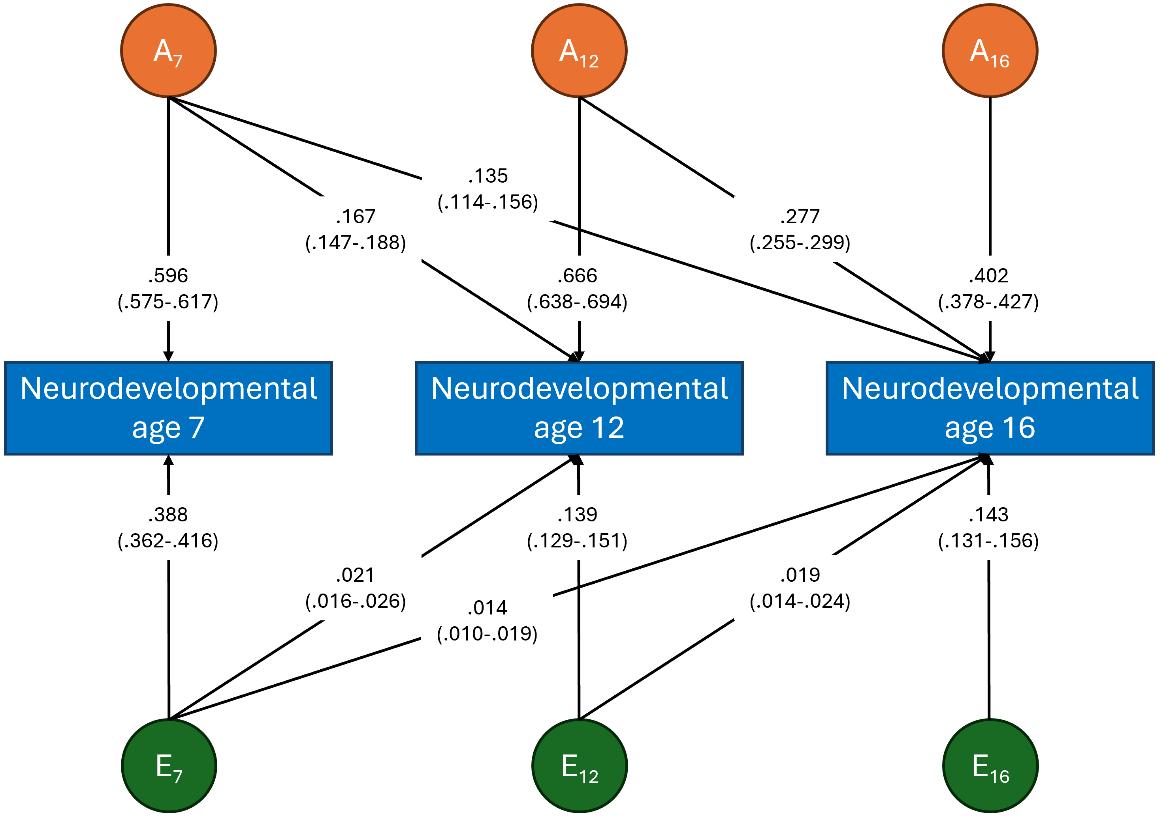

**Supplementary Figure 2**. Results of the Cholesky decomposition of the covariation between neurodevelopmental factors across time points.

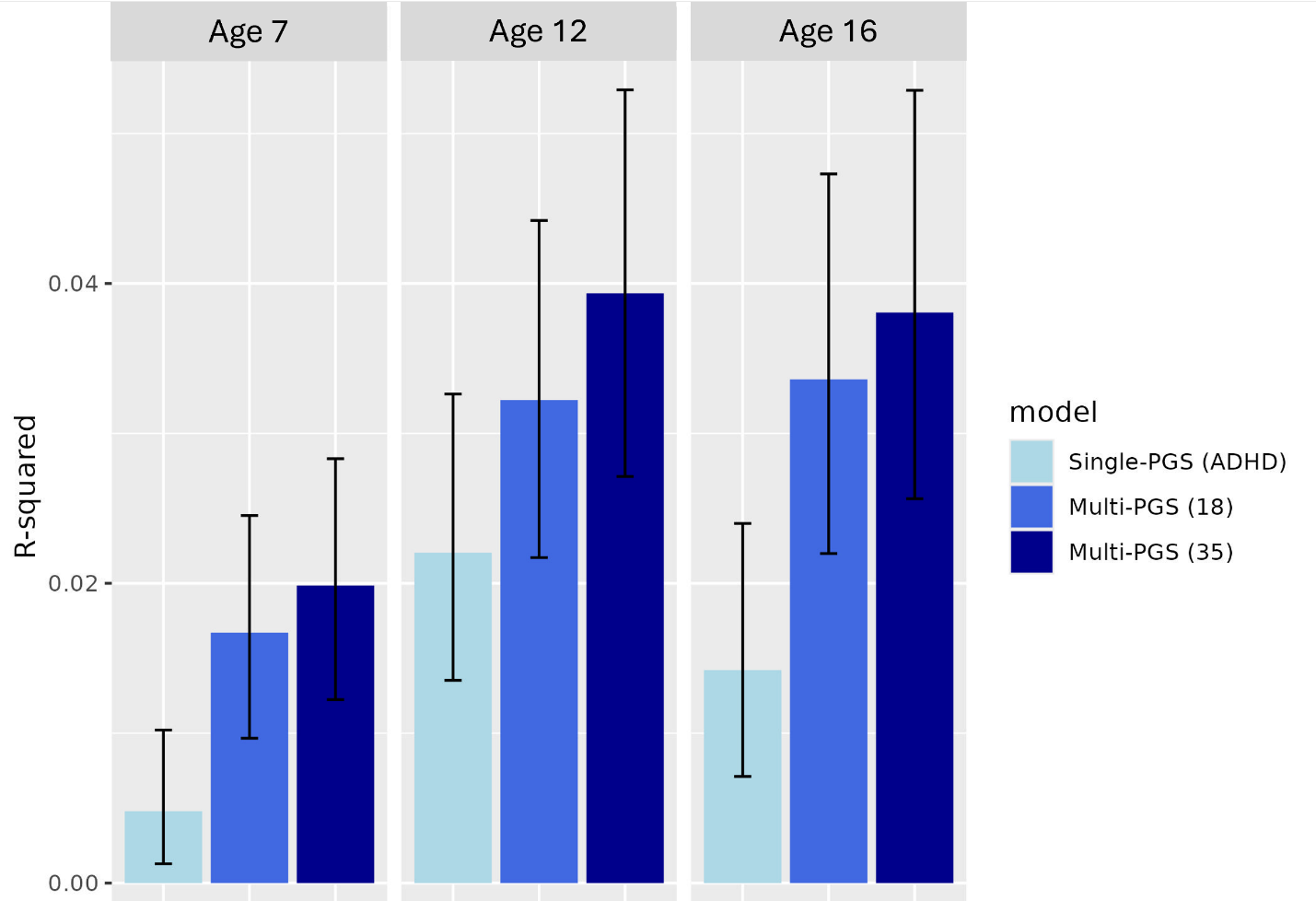

**Supplementary Figure 3**. Variance explained (R-squared) in neurodevelopmental factors at age 7, 12 and 16 by the most predictive polygenic score (PGS), i.e., ADHD, jointly by the 18 PGS included in primary analyses, and jointly by a wider set of 35 PGS. For a full list of PGS included in each model see Supplementary Tables 2 and 13.
